# Wastewater Sequencing Reveals Persistent Circulation and Rising Prevalence of Several Oncogenic Viruses Across Texas

**DOI:** 10.1101/2025.09.17.25335998

**Authors:** Harihara Prakash, Ryan K. Perez, Matt Ross, Michael Tisza, Sara J. Javornik Cregeen, Jennifer Deegan, Joseph F. Petrosino, Eric Boerwinkle, Justin R. Clark, Anthony W. Maresso

**Author notes:** **Correspondence:** Anthony W. Maresso, Ph.D, Justin R. Clark, Ph.D, Eric Boerwinkle, Ph.D.

## Abstract

**Background:** Oncogenic viruses cause high-risk cancers in humans and are responsible for nearly 20% of all cancer cases worldwide. Currently, very limited data exists in the realm of wastewater-based viral epidemiology (WBE) of cancer-causing viruses, with existing studies using targeted approaches (i.e PCR-based approaches) which lack scalability. Our study aims to carry out WBE with hybrid-capture probes to detect and track multiple oncogenic viruses simultaneously in wastewater across Texas, USA, overcoming the drawbacks associated with targeted approaches.

**Methods:** Here, we used a hybrid-capture approach to detect, filter and sequence oncogenic virus signals from wastewater samples collected over a duration of three years, from May 2022 to May 2025. Once viral reads were sequenced, we utilized established computational tools to characterize reads into their respective virus of origin. Next, viral abundances of each characterized oncogenic virus were tracked over time and read coverage across their genomes was measured using read mapping techniques.

**Findings:** We detected six known oncogenic viruses, along with three suspected oncogenic viruses across all sampling locations within Texas. Over three years, viral abundance gradually increased, with distinct peaks and dips over the summer and winter months. The prevalence of high-risk viruses such as HPV and EBV rose sharply, with increases in abundance observed post-2024. We also obtained nearly 100% genome coverage with viral reads captured using a hybrid-capture technique for almost all oncogenic viruses and their types.]

**Interpretations:** Our study shows that a hybrid-capture method can efficiently overcome the challenges faced with using targeted approaches for WBE. Using this method, we get broader read coverage, coupled with concurrent and consistent real-time tracking dynamics of multiple oncogenic viruses. Our findings also emphasize the persistent circulation and rising prevalence of high-risk cancer-causing viruses, underscoring the need for sustained public health interventions to protect communities and assess viral prevalence in high-risk populations.

**Funding:** This work was supported by S.B. 1780, 87th Legislature, 2021 Reg. Sess. (Texas 2021), the Baylor College of Medicine and the Alkek Foundation Seed Funds.

**Research in context:** *Evidence before this study:* Cancer-causing viruses are of major clinical significance, responsible for nearly 20% of all recorded cancer incidences in humans worldwide. With some of these viruses causing high-risk cancers such as cervical cancer, there is a need for improved detection, tracking and control of oncogenic viruses across the globe. During the SARS-CoV-2 pandemic, wastewater-based viral epidemiology (WBE) rose to the forefront of virus tracking, utilizing non-invasive methods to detect and monitor the prevalence of medically relevant viruses of concern. Today, WBE has been utilized to accelerate the surveillance of numerous viruses such as SARS-CoV-2, mpox, influenza and more. To reinforce our prior knowledge on current trends in WBE of oncogenic viruses, we searched for research articles in PubMed and Google Scholar containing keywords “wastewater” and “oncogenic viruses” or “tumor viruses”. Further, to look specifically at oncogenic viruses of clinical concern, we searched for studies containing the keywords “wastewater” along with each of the 9 oncogenic viruses—Human papillomavirus, Hepatitis B virus, Hepatitis C virus, BK virus, Epstein-Barr virus, Merkel cell polyomavirus, Human polyomavirus, Kaposi Sarcoma-associated virus and Human T-cell lymphotropic virus type 1. On assessment of the search results, we found that specific oncogenic viruses had been detected in the wastewater of numerous countries including Egypt, Uruguay, Canada, Catalonia, Italy, India and Australia. These studies primarily focused on using PCR-based techniques to sequence and obtain viral read sequences from wastewater, measuring viral RNA concentrations in the process. While most of these studies focused on sole detection of oncogenic viruses, some included the analysis of prevalence of viral load over time. However, we found no other studies showcasing concurrent tracking dynamics of multiple oncogenic viruses in the United States over a span of three years.

*Added value of this study:* Considering the medical significance of oncogenic viruses, we set out to utilize Texas Wastewater and Environmental Biomonitoring’s (TexWEB’s) hybrid-capture approach to detect and track these viruses in wastewater. Current WBE approaches utilize PCR-based techniques, which have drawbacks such as limited specificity to a single target. Our study highlights the value in using a hybrid-capture method to bring forth a greatly improved WBE approach, providing near real-time tracking of all oncogenic viruses. To our knowledge, our work is the first comprehensive WBE approach which uses a sequencing-based method to detect all known oncogenic viruses concurrently. Here, we present the tracking of viral abundances over the course of three years for all viruses, analyzing seasonal variations in the process. Further, we also showcase the ability to identify genomic regions on viral reference genomes from which sequenced reads originate. This information can be an invaluable tool towards understanding the dynamics of the prevalence of cancer-causing viruses in the general population, their relationship to cancer incidences in humans, and their mechanisms of viral evolution.

*Implications of all the available evidence:* Our study highlights the advantages of using a hybrid-capture approach for WBE of oncogenic viruses. This approach can be used to detect a complete panel of multiple selected viruses and provides viral abundance and prevalence information, overcoming the drawback of high specificity to single targets that come with PCR-based approaches. We also highlight the scope of this approach in tracking and monitoring multiple relevant oncogenic viruses concurrently at regular sampling intervals. Data generated using this technique can be used by public health departments to identify viruses of immediate concern, set up pandemic preparedness and intervention programs, as well as promote vaccination drives while spreading general public health awareness among communities.

## 1. Introduction

It is estimated that oncogenic viruses are responsible for 20% of all human cancers.^1^ These viruses span at least six distinct viral families^2^ and can drive cancer formation through a variety of diverse mechanisms, including insertional mutagenesis, integration into the host genome, or inhibition of tumor suppressor pathways.^2^ They have been linked to cancers of the cervix, anus, vagina, oropharynx, liver and other sites, with broad clinical implications.^3,4^ Among the known oncogenic viruses are high-risk Human papillomaviruses (HPV), Hepatitis B virus (HBV), Hepatitis C virus (HCV), Epstein-Barr virus (EBV), Human T-cell lymphotropic virus (HTLV-1), and Kaposi sarcoma herpesvirus (KSHV). These viruses are classified as Group 1 carcinogens by the International Agency for Research on Cancer. Others, like Merkel cell polyoma virus (MCPyV) and BK polyomavirus (BKV), and other Human polyomaviruses (HPyV) are strongly linked to cancer and known carcinogens.^5^

Because these viruses often establish chronic, persistent infections, symptoms may not appear until years or decades after exposure. Cervical cancer caused by high-risk HPV, for example, develops slowly over time, while hepatocellular carcinoma (HCC) resulting from chronic HBV infections is not seen until middle age or later.^6,7^ Thislong latency period complicates public health efforts to interrupt transmission and intervene before cancer develops.

Wastewater-based viral epidemiology (WBE) is a promising public health surveillance approach to monitor viral infections at the population level.^8^ WBE has emerged as a valuable, non-invasive tool during the SARS-CoV-2 pandemic. Traditionally, WBE employs bioanalytical methods such as liquid chromatography-tandem mass spectrometry (LC-MS/MS) and quantitative polymerase chain reaction (qPCR), letting researchers track the abundance of medically important biomarkers in wastewater.^9,10^ Detection of biomarkers excreted into sewage at the community level allows for the assessment of the overall health of a population and cross-analysis with clinical cases.

The Texas Epidemic Public Health Institute (TEPHI) launched the TexWeb initiative in May 2022 to track viral outbreaks using a broader approach.^11^ Instead of targeting individual pathogens, TexWEB uses a custom hybrid-capture sequencing approach using comprehensive viral probes designed by Twist Biosciences. This probe set is designed to enrich for over 3,000 human and animal viruses, including high-impact viruses such as mpox, SARS-CoV-2, influenza, dengue and more.^8,12-15^ As of 2025, this system routinely samples and analyzes wastewater from more than 40 sites in 15 cities across Texas, monitoring over 500 different viruses and reporting over 15,000 different viral variants.^11^

In this study, we apply this hybrid-capture technique to track the temporal distribution patterns of known oncogenic viruses. While all known oncogenic viruses were detected by the probe set, we focus here on three: HBV, HCV, and HPV. Although Hepatitis A virus (HAV) and Hepatitis E virus (HEV) are not oncogenic, we briefly include them to provide context for interpreting hepatitis viral dynamics more broadly.^16^ We also acknowledge the association Hepatitis D virus (HDV) has with HBV in promoting HCC in afflicted individuals, and have included a closer look at this virus as well.

HBV and HCV are the primary causative agents of HCC, responsible for an overwhelming 80% of all cases.^3^ The incidence of HCC is predicted to have an estimated incidence of more than one million cases by the end of 2025, driven by chronic hepatitis infections that progress to liver cirrhosis and liver failure, and eventually cancer and death.^17^ HDV while not oncogenic in itself, can still co-infect a host with HBV, accelerating the development of HCC by increasing the rate of progression towards life-threatening liver cirrhosis.^25,26^

HPV, meanwhile, causes nearly 95% of all cervical cancer in women worldwide, with over 42 million infections recorded in 2018 in the United States alone.^4,18^ Among the nearly 200 known genotypes, HPV16 and 18— members of the alpha-papillomavirus genus—are responsible over 70% of all recorded cervical cancer cases.^4,19,20^ HPV genera are classified into alpha-papillomavirus, beta-papillomavirus, gamma-papillomavirus, mu-papillomavirus and nu-papillomavirus based on the sequence of the open reading frame coding for the L1 capsid protein.^21^

Here, we demonstrate that hybrid-capture sequencing enables robust detection and genomic characterization of oncogenic viruses in wastewater across Texas. Our results show high read depth and broad genome coverage for key pathogens, including the ability to track HPV diversity at the genus level. These findings support the use of WBE as a surveillance tool for cancer-associated viruses and lay the groundwork for future efforts that link wastewater signals to clinical outcomes and targeted interventions.

## 2. Results

### Oncogenic viruses are consistently detected in wastewater across Texas

To evaluate the presence of oncogenic viruses in wastewater samples and validate the utility of hybrid-capture sequencing, we analyzed samples collected by TexWeb over a three-year period (May 2022 to May 2025). Oncogenic viruses were consistently detected across this time frame, with ongoing monitoring at urban catchment sites throughout the state.

When viral read counts were aggregated across all sampling sites, hybrid-capture sequencing recovered signals for all nine oncogenic viruses. Average RPKMF values varied across viruses and over time, with HPV, MCPyV, EBV and BKV showing a gradual upward trend in viral abundance over the three-year period (**Fig. 1**). This data supports the feasibility of hybrid-capture as a consistent and sensitive method for detecting oncogenic viruses in complex environmental samples.

**Fig. 1:**
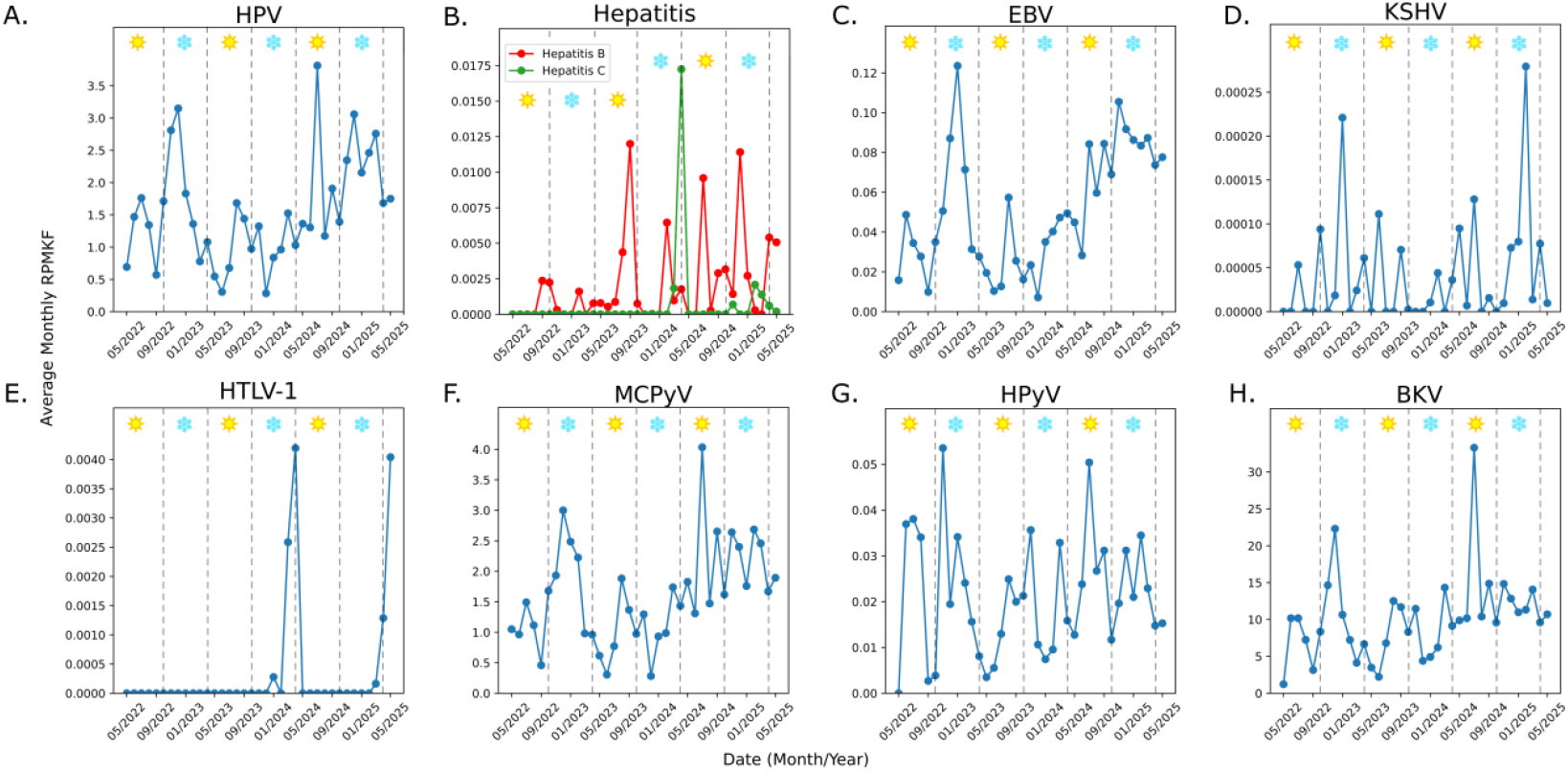
(A-H): Abundance of Oncogenic Virus Reads over Three Years of Sampling Across Texas. Mean viral signal measured in average monthly RPKMF over time in samples taken from wastewater and measured using hybrid-capture probes. Warmer months are highlighted with a sun icon (April-September). Colder months are highlighted with a snowflake icon (October-March). Seasonal boundaries were defined using statewide average temperatures to determine the midpoints between the summer and winter temperature extremes. Both oncogenic and non-oncogenic HPV types are included as one in **Fig. 1A**. Acronyms: HPV, Human papillomavirus; EBV, Epstein-Barr virus; HTLV-1, Human T-cell lymphotropic virus type 1; KSHV, Kaposi sarcoma herpesvirus; MCPyV, Merkel cell polyomavirus; HPyV, Human polyomavirus (Specifically Alpha- and Delta-polyomavirus); BKV, BK polyomavirus.

Some viruses, such as HPV (both oncogenic and non-oncogenic types), EBV, MCPyV, and BKV (**Fig. 1A, 1C, 1F, and 1H**), were detected in at least one site nearly every month, indicating stable and widespread presence. In contrast, others, including Hepatitis C, KSHV and HTLV-1 (**Fig. 1B, 1D, and 1E**) exhibited more frequent non-detection or low read abundance, with RPKMF values rarely exceeding 0.02. Despite this, these low-abundance viruses were still detected intermittently across multiple sites, demonstrating the method’s capacity to capture even sparse signals. The prevalence of HPyV, alpha- and delta-polyomaviruses taken together in particular (**Fig. 1G**), was found to show no distinct seasonal pattern, with dips and peaks throughout the sampling duration.

Among the Hepatitis viruses, HBV and HCV were detected in low amounts (**Fig. 1B**), with HCV detections very sparse throughout the entire sampling duration with low prevalence observed across all sites. To get a full picture of Hepatitis transmission dynamics across Texas over three years of sampling, we looked at the abundance of all Hepatitis viruses together (**Supp. Fig. 1**). We observed sparse detections for HDV and HEV, while the prevalence of HAV was seen in the form of sporadic peaks and dips across the complete sampling duration. Taken together, these patterns indicate that hybrid-capture sequencing can successfully recover both high-abundance and rare oncogenic viruses in wastewater and supports its use for long-term surveillance.

Building on these observations, we next analyzed genome coverage for six representative viruses with high clinical significance—Hepatitis A, Hepatitis B, Hepatitis C, Hepatitis D, Hepatitis E and HPV—to assess read depth and completeness of genome recovery.

### Hybrid-capture sequencing recovers broad genomic coverage from wastewater

Considering the medical relevance of the Hepatitis virus family to the healthcare community, we chose to take a deeper look at all Hepatitis viruses—both oncogenic and non-oncogenic—to establish a solid basis for comparison. To evaluate genomic breadth and depth of coverage, we performed detailed mapping of reads assigned to all Hepatitis viruses (Hepatitis A, Hepatitis B, Hepatitis C, Hepatitis D and Hepatitis E) using a best-to-all alignment strategy. Reads classified as belonging to a given virus were mapped to all reference genomes for that virus included in the EsViritu Virus Pathogen Database.^23^ The reference accession receiving the highest number of mapped reads was selected, and all matching reads were realigned to that reference for visualization and coverage analysis.

For HAV, 93,846 reads mapped to the selected reference genome (Accession: LC373510), with coverage spanning nearly the entire genome at high depth. Read density was particularly strong across the VP1, VP2, and VP3 regions, but the signal extended into other coding and non-coding regions as well (**Fig. 2A**).

**Fig. 2:**
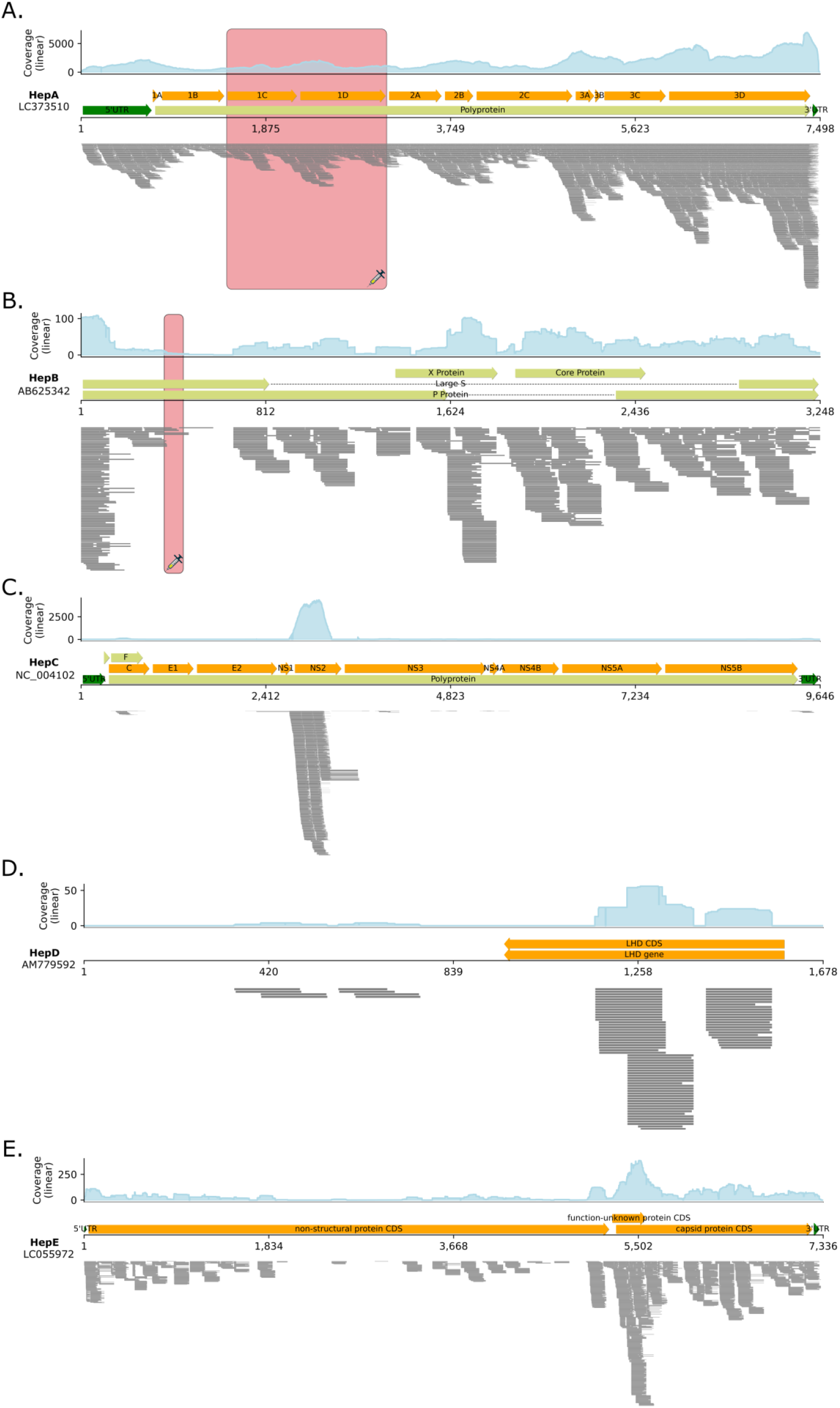
Genome-Wide Read Coverage of Sampled Hepatitis Viruses via Hybrid-Capture Sequencing. Histogram **(top)** shows coverage depth across complete genome. Genomic annotations across the genome are highlighted with arrows **(middle)**. Read distribution shown by stacked reads **(bottom)**. Vaccine target regions on the relevant Hepatitis genomes are highlighted in red with a syringe icon. **(A)** Reads mapped to reference Hepatitis A genome (Accession: LC373510). **(B)** Reads mapped to reference Hepatitis B genome (Accession: AB625342). **(C)** Reads mapped to reference Hepatitis C genome (Accession: NC_004102). **(D)** Reads mapped to reference Hepatitis D genome (Accession: AM779592). **(E)** Reads mapped to reference Hepatitis E genome (Accession: LC055972).

In contrast, relatively fewer reads were recovered for HBV. Of the 771 classified reads, 765 mapped to the selected reference genome (Accession: AB625342), with coverage observed across multiple functional regions. Despite the lower total count, reads aligned consistently to key genes, including the S (surface) gene, P (polymerase) gene, C (core) gene and X gene regions, suggesting that meaningful genomic information can still be recovered from low-abundance targets (**Fig. 2B**).

In the case of HCV however, 10,728 out of 10,862 classified reads were mapped to the chosen reference genome (Accession: NC_004102), but coverage was more fragmented. Reads primarily aligned to the core protein, NS2, and NS3 regions, with a disproportionately large number of the mapped reads mapping specifically to the NS2 region. Full genome coverage was not achieved, though multiple coding regions were well represented (**Fig. 2C**). To test for the possibility of expression vector contamination, we randomly selected mapped reads, and looked at their top BLAST (Basic Local Alignment Search Tool) hits from the NCBI database.^27,28^ We found that the number of non-synthetic HCV hits greatly outnumbered the synthetic hits, indicating vector contamination is unlikely in the case of HCV. These results confirm that hybrid-capture sequencing enables broad genome recovery for both high- and low-abundance viral targets, and can provide valuable sequence information even when total read counts are limited.

We observed that the least reads recovered from the wastewater belonged to HDV. Of the meager 96 HDV reads obtained, 88 reads were found to map to the reference genome (Accession: AM779592). Interestingly, coverage of these reads was mostly limited to the LHD gene region, with over 90% of mapped reads mapped to this genomic region **(Fig. 2D)**.

Finally, of the 2,847 HEV reads sampled from wastewater, 2,341 reads mapped to the selected HEV reference genome (Accession: LC055972). While reads mapped sparsely to regions throughout the reference genome, a distinctly large number of reads mapped to the capsid protein CDS of the HEV genome, indicating shedding of this protein in relatively larger levels into the wastewater **(Fig. 2E)**.

### HPV genera show distinct prevalence patterns in wastewater

We assigned reads classified as HPV to one of five genera: alpha-, beta-, gamma-, mu- and nu-papillomaviruses based on taxonomic labels provided by EsViritu. During the three-year sampling period, beta-HPV consistently showed the highest average abundance statewide, followed by gamma-HPV (**Fig. 3A**). These two genera also exhibited reoccurring semi-seasonal twice-yearly spikes in read abundance, most notably in mid-2022 (May-July), late 2022 (September-December), mid-2023 (June-August), and mid to late 2024 (June-December). Alpha-HPV, including the high-risk type 16 and type 18 strains, on the other hand, was present in lower overall levels but demonstrated short-lived peaks that aligned with most of the broader multi-genus spikes, including those in July 2022, August 2023, and July 2024.

**Fig. 3:**
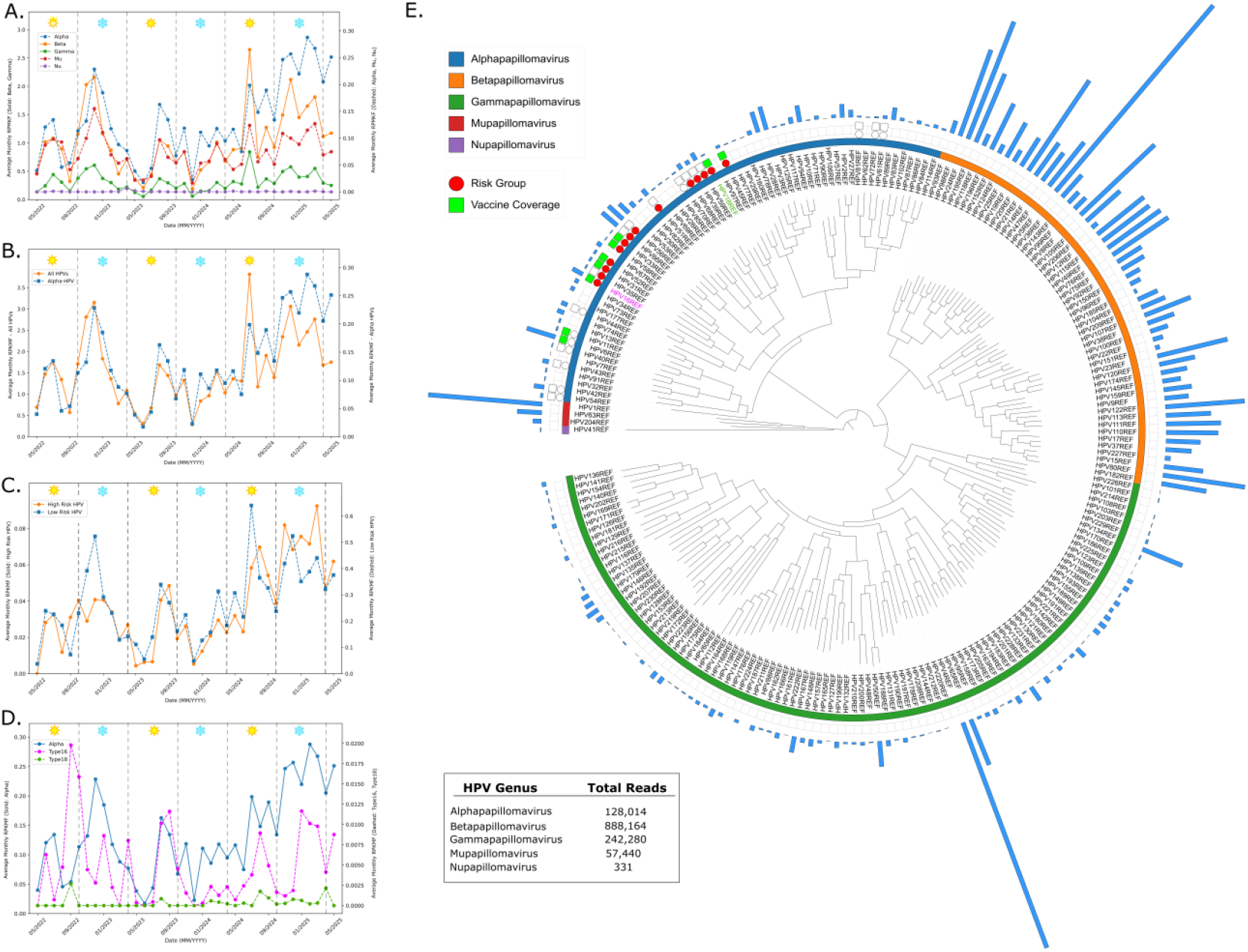
HPV Genera-Specific Abundance and Phylogenetic Resolution of Wastewater-Derived Reads. (**A**) Prevalence of alpha-, beta-, gamma-, mu- and nu-HPV genera measured as average RPKMF from May 2022 to May 2025. (**B**) Comparison of highly oncogenic alpha-HPV against all other HPV genera. **(C)** Comparison of high-risk HPV against low-risk HPV **(D)** Comparison of alpha-HPV type 16 and alpha-HPV type 18 against all alpha-HPV types. **(E)** Phylogenetic reconstruction of HPV genera and their types. Outer bar chart represents number of reads mapping to each type. Risk grouping of high-risk alpha-HPV types are highlighted with a red circle, while vaccine coverage is highlighted with a green square. Empty circles and squares indicate low-risk types and zero vaccine coverage respectively. High-risk alpha-HPV type 16 and alpha-HPV type 18 labels are highlighted in pink and green respectively. Warmer months are highlighted with a sun icon (May-September). Colder months are highlighted with a snowflake icon (October-April).

Mu- and nu-HPV were detected at very low levels throughout the study period, with nu-HPV appearing only sporadically. These genera contributed minimally to the overall HPV signal.

An interesting extended peak in alpha-HPV was observed in early 2025, which followed a multi-genus increase in late 2024. This suggests that while alpha-HPV abundance often tracks with large HPV subfamily trends, it may occasionally follow a different pattern. These genus-level differences may reflect underlying biological distinctions: alpha-HPVs are typically associated with mucosal surfaces, while beta- and gamma-HPVs are associated with cutaneous infections and a broader environmental presence.

When comparing alpha-HPV specifically to the total HPV signal across all genera, we found that two trends rose and fell in parallel overall (**Fig. 3B**). However, alpha-HPV remained consistently lower in abundance and showed distinct peak timing in February 2025, where it peaked roughly two months after the winter spike in total HPV signal, suggesting that alpha-HPV may exhibit slightly different dynamics in wastewater than other genera.

### HPV wastewater reads resolve into clinically relevant phylogenetic lineages

To evaluate the diversity and taxonomic resolution of HPV sequences detected in wastewater, we built a maximum-likelihood phylogenic tree using representative HPV genomes from the PaVE (The Papillomavirus Episteme) database.^22^ Each branch tip was colored by genus and bar charts were overlaid showing how many reads mapped to each genome (**Fig. 3E**). HPV genomes were also annotated with symbols for risk classification (high-risk and low-risk) and vaccine coverage.

Mapping data confirmed broad representation across alpha-, beta-, and gamma-papillomaviruses. Beta-HPV types showed the widest spread, with low to moderate read support across a broad range of genomes. One genome, beta-HPV type 5 (HPV5REF), dominated within this genus, receiving over 95,000 mapped reads. Similarly, in the gamma genus, gamma-HPV type 4 (HPV4REF) had the strongest support with over 93,000 reads.

In contrast, alpha-HPV types, which include high-risk mucosal genotypes, showed tighter clustering of mapped reads. Among the alpha-HPV genotypes detected, alpha-HPV type 6 (HPV6REF) and alpha-HPV type 3 (HPV3REF) accounted for the highest number of mapped reads. Both are considered low-risk types associated with benign mucosal infections. We also recovered several high-risk, cancer-associated genotypes, including alpha-HPV type 16 (HPV16REF) and alpha-HPV type 18 (HPV18REF), the two genotypes responsible for most HPV-associated cancers. These oncogenic types were consistently present across the time series despite their lower abundance (**Fig. 3D**).

In total, all nine HPV types targeted by the 9-valent vaccine (HPV6, 11, 16, 18, 31, 33, 45, 52 and 58) were identified in our phylogenetic tree. This demonstrates that the hybrid-capture sequencing approach can detect both high-risk and low-risk HPV types from complex environmental samples, including vaccine-preventable lineages (**Fig. 3C**).

This approach confirmed representation across alpha-, beta-, and gamma-papillomaviruses with mapped reads spanning much of the phylogenetic tree. Alpha-HPV genotypes, including high-risk cancer-associated types, were reliably recovered which supports the ability of hybrid-capture sequencing to resolve clinically relevant alpha-HPV types.

### HPV genomes achieve near-complete coverage with hybrid capture sequencing

To visualize genome-wide coverage within each HPV genus, we aligned all available genomes per genus using MAFFT and selected the genome within the genus with the highest read count to act as an anchor. Reads from all members of a given genus were then transposed to this anchor genome coordinate space to generate read stack plots and coverage maps, allowing for intra- and inter-genus comparisons of sequencing depth and coverage breadth (**Fig. 4**).

**Fig. 4:**
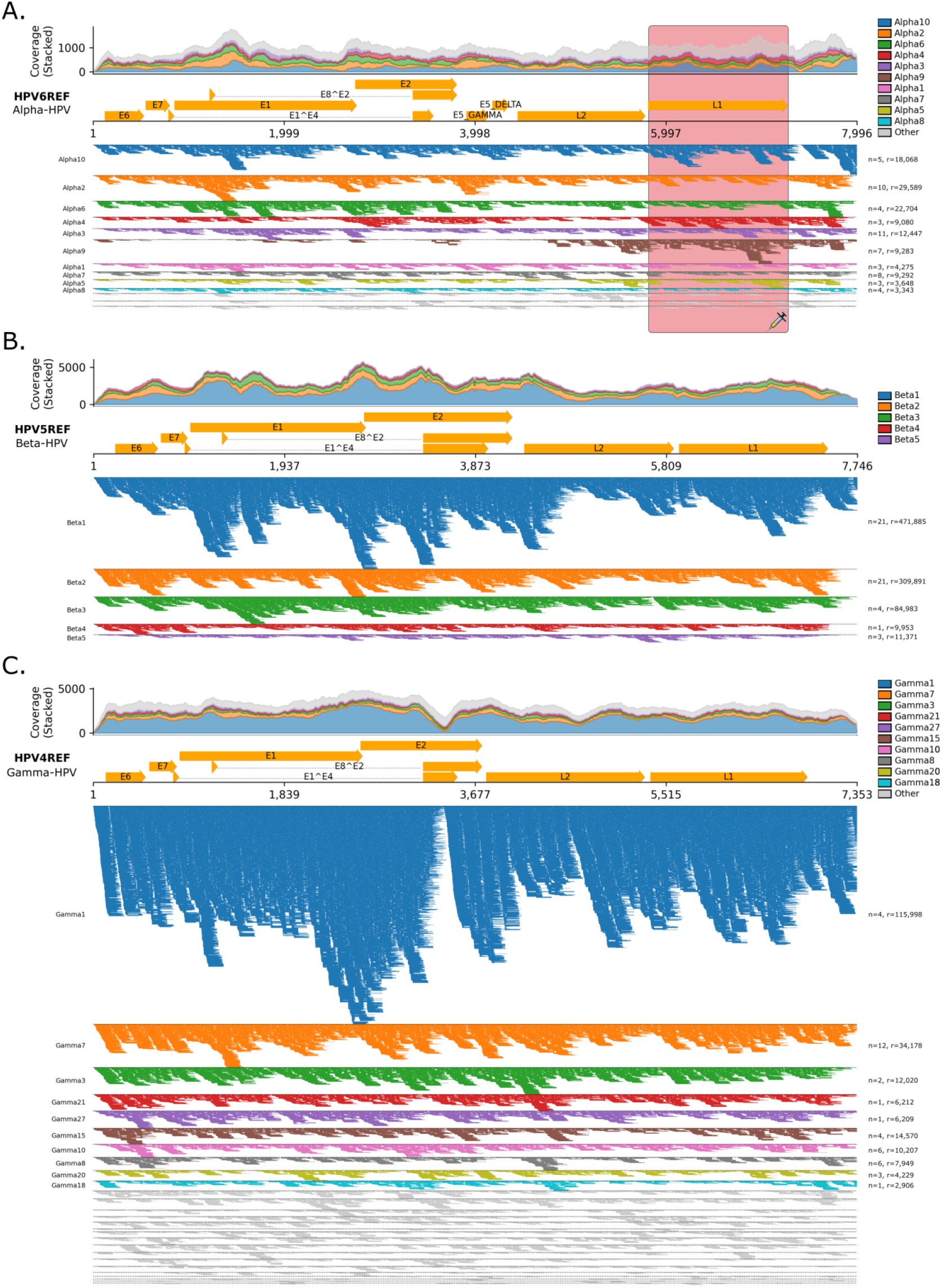
Genome-Wide Coverage of HPV Genera from Wastewater Using Hybrid-Capture Sequencing. Multi-stack coverage maps of alpha-, beta- and mu-HPV reads mapped to their respective genus-level species types. Top 10 species **(left)** highlighted with non-gray colors while the remaining species are highlighted in gray. Stacked coverage map **(top)** showcases coverage of reads mapped to top 10 species. Number of types per genus-level species (n) and number of reads mapped (r) are also shown **(right)**. Vaccine target regions on the genome highlighted in red with a syringe icon. (**A**) Alpha-HPV Reads mapped to all alpha-HPV species. (**B**) Beta-HPV Reads mapped to all beta-HPV species. **(C)** Mu-HPV Reads mapped to all Mu-HPV species.

In **Fig. 4A**, we show the alpha-HPV coverage map, with reads transposed onto HPV type 6 (HPV6REF). Read stacks were primarily concentrated around two low-risk mucosal types, HPV type 6 and HPV type 3 (HPV3REF), which together accounted for most alpha-HPV-classified reads. Coverage across the genome was variable, but key regions such as L1, L2, and E6/7 oncogenes showed consistent signal, confirming that hybrid-capture sequencing recovers clinically relevant alpha-HPV regions from wastewater.

**Fig. 4B** shows beta-papillomavirus coverage using HPV type 5 (HPV5REF) as the anchor genome. This genus had the broadest read support across types, with HPV type 5 contributing the largest fraction. Coverage was even across most of the genome, indicating widespread environmental presence and consistent detection of beta-HPV types.

In **Fig. 4C**, gamma-HPV reads were transposed to HPV type 4 (HPV4REF). Like beta, the gamma-HPV signal was dominated by a single high-abundance type but also showed higher baseline support with broad and relatively uniform coverage across all types.

## 3. Discussion

Our work showcases the ability to detect and track all known oncogenic viruses in wastewater samples across vast geographic regions using a sequencing-based approach. Building on previous WBE studies, we demonstrate the utility of a hybrid-capture technique to detect oncogenic viruses from wastewater, classifying and characterizing them at the genus level. Using similar methods previously applied to track viruses such as mpox and avian influenza^13-15^, we extended our approach to viruses known to cause cancer, including HPV, Hepatitis viruses, BKV, and others. Over three years of sequencing, we characterized nearly 9,000,000 viral reads into nine distinct oncogenic viruses, as well as all members of the Hepatitis viral family, measuring their abundance and identifying genomic regions from which the reads originated. By combining changes in viral signal over time with changes within the viral genome, we can track current clinically relevant strains and substrains, while maintaining an overview of all oncogenic viruses across the geographic landscape. Such information is valuable for risk assessment and for developing public health policies that inform officials about the degree of risk in their communities. Tracking the evolution of high-risk viral strains at the molecular level can also guide the design of new vaccines, identification of novel drug targets, and development of diagnostic tools.

After 2024, we observed peak abundance for several oncogenic viruses, including HTLV-1, HPV, HCV, KSHV, MCV, and BKV. Notably, viral loads increased sharply across all targets during the summer months of 2023 and again in 2024. While no consistent annual seasonal cycle was evident, these summer spikes may reflect increased travel and interpersonal contact during vacation and holiday periods in academic settings such as colleges. The removal of pandemic-related social distancing could also have contributed to higher transmission of sexually transmitted oncogenic viruses such as HPV, and likely underlies the sharp spike observed in late 2022 that coincided with post-COVID mixing and the immunity debt period documented for respiratory viruses such as the Respiratory Syncytial Virus (RSV).^24^ Some viruses, including HTLV-1 and KSHV, were detected at disproportionately low levels relative to others, which may reflect low prevalence in wastewater or reduced capture efficiency of the hybrid-capture probes.

Once we classified viral reads into their respective hepatitis types, we examined genome coverage across reference genomes. Near-complete coverage was achieved for HAV and HBV, with semi-complete coverage obtained for HEV. This demonstrates the hybrid-capture approach’s ability to recover full viral genomes from wastewater. In contrast, HCV and HDV coverage was highly fragmented, with nearly 95% of reads mapping to a short segment in the NS1/NS2 region for HCV, and almost 90% of reads mapping to segments in the LHD region for HDV. Multiple HAV reads and a smaller number of HBV reads mapped to vaccine target regions, indicating that this method can recover sequences relevant for monitoring potential resistance mutations or immune escape.

To further assess the taxonomic resolution, we focused on HPV due to its medical relevance, multiple genera, and varying health risks. Across three years, beta- and gamma-HPV showed higher abundance than alpha-, mu-, and nu-HPV, likely reflecting more efficient cutaneous transmission of beta and gamma types compared to sexual transmission routes of alpha types. Although less abundant, alpha-HPVs are clinically significant because many types are high-risk mucosal genotypes linked to cancer. Nu-HPV was rarely detected, with its highest levels appearing in late winter 2023.

Comparing alpha-HPVs to all genera showed lower overall abundance but similar timing of peaks and dips. This pattern suggests that HPV prevalence across sites is not driven by a single genus, with abundance in multiple genera tending to rise and fall together. Monitoring these parallel trends could help identify broader shifts in HPV transmission within the community.

When comparing high-risk and low-risk alpha-HPV types, high-risk variants were consistently less abundant. This pattern may reflect the impact of vaccination campaigns targeting high-risk types, which could lower their prevalence in wastewater. In contrast, low-risk types, which are not typically vaccine targets and pose less severe health risks, were detected in higher abundances across the state (**Fig. 3C**).

HPV16 and HPV18 together cause more than 70% of cervical cancer, yet wastewater data showed HPV16 in higher abundance than HPV18. HPV16 trends generally mirrored those of all alpha-HPV types, whereas HPV18 remained low except for small peaks in summer months. These differences may reflect variation in shedding, persistence, transmissibility, or detection sensitivity. Correlating wastewater trends with clinical data will be important to determine whether the lower HPV18 signal reflects true prevalence or under-detection.

Finally, we assessed coverage depth and breadth for HPV at the genus and type levels. Mapping showed reads across multiple types within each genus, with near-complete genomic coverage for most. The major exception was nu-HPV, where coverage was sparse due to low abundance. These results confirm that the hybrid-capture approach can resolve HPV diversity to its types, enabling independent tracking of each lineage.

Although hybrid-capture recovered near-complete genomes for many oncogenic viruses, coverage varied across targets. HCV reads were concentrated in the NS1/NS2 region, HDV reads were found in the LHD region, and nu-HPV coverage was sparse reflecting both low abundance and possible probe bias. These observations suggest that genome recovery from wastewater may depend on factors such as viral persistence, transmissibility, and shedding dynamics, and that not all oncogenic viruses are equally amenable to broad genome reconstruction with this method.

Future directions should integrate wastewater signals with vaccination data to evaluate how immunization campaigns influence the prevalence of high-risk viruses such as HBV and HPV. This approach could help test whether vaccination reduces oncogenic virus abundance in the community. In addition, variant analysis may provide insight into the evolution of high-risk strains and allow for early identification of mutations that enhance viral fitness or reduce vaccine efficacy.

In conclusion, this study demonstrates that sequencing enables consistent detection and genomic characterization of oncogenic viruses in wastewater across large geographic regions. The approach provides sufficient resolution to monitor viral genera and subtype, including high-risk lineages, and to assess genomic variation relevant to transmission and resistance. As wastewater-based epidemiology expands, these findings highlight the potential of sequencing-based methods to support long-term surveillance of cancer-associated viruses and inform public health strategies.

## 4. Methods

### Sample collection, shipping and processing

Wastewater samples from over 40 sites across 15 cities in Texas were collected, shipped and processed using methods described by Tisza *et al*. 2024.^14,22^ 100-500 mL of raw wastewater was collected and stored in 500 mL prelabeled sample bottles. Collection sites were coded upon the request of public health officials to mask the identity of the cities where sampling was carried out.

To decontaminate the surface of the collection bottles, 10% bleach was used followed by sealing of samples into biohazard bags within shipping boxes containing ice packs and absorbent pads. The boxes were then shipped to the Alkek Centre for Metagenomics and Microbiome Research at the Baylor College of Medicine, Houston, Texas. Upon arrival, samples were barcoded and stored at 4 °C prior to processing.

Samples were processed as described above, with the objective of extracting bulk DNA and RNA from centrifuged solid pellets.^22^ Reverse-transcription of RNA to cDNA is first carried out with the Protoscript II First Strand cDNA Synthesis Kit (New England Biolabs Inc.), NEBNext Ultra II Non-Directional RNA Second Strand Module (New England Biolabs Inc.) and Random Primer 6 (New England Biolabs Inc.) before mixing it with DNA for library construction. Library construction is carried out with Twist Library Preparation EF 2.0 Kit and Twist Universal Adaptor System (Twist Biosciences) and pooled with 16 samples, totaling 1,500ng. Hybridization of probes was carried out with the Twist Comprehensive Viral Research Panel (Twist Biosciences) at 70 C for 16 h. Libraries were sequenced on Illumina NovaSeq 6000 SP flow cells, producing raw data files in the binary base call (BCL) format. The raw BCL files were converted to FASTQ format and demultiplexed based on the dual-index barcodes using the Illumina ‘bcl2fastq’ software.

### Oncogenic virus sequence assignment with EsViritu

To assign sequences to their corresponding virus, the demultiplexed FASTQ files are first processed with BBDuk for quality trimming (Q25). Adapter trimming and removal of reads mapping to PhiX Illumina spike-in is carried out with BBMap. Reads mapping to the human reference genome (GCF_000001405.39) are also filtered out. Next, the remaining reads are classified using EsViritu (v0.2.3), a previously published bioinformatics pipeline.^23^ Here, the processed reads are aligned to the complete Virus Pathogen Database (v2.0.2)^23^ using minimap2, keeping alignments with at least 90% average identity across 90% of the read length.

At low levels in wastewater, the proportion of reads aligning to virus genomes is presumed to correspond to the proportion of viral nucleic acid molecules over the remaining molecules in the sample.^23^ Thus, we used RPKMF (calculated as Reads Per Kilobase of reference genome/Million reads passing Filtering) as our abundance metric over the conventionally used RPKM (calculated as Reads Per Kilobase of reference genome/Million reads mapped to reference sequences) metric. With these RPMKF values, the per sample abundances, coverage metrics and taxonomic information were tabulated into a single sheet and used for downstream analysis.

### Taxonomic filtering and abundance estimation using TREx

Before analyzing oncogenic virus signals from the EsViritu output, all accession ID’s of known oncogenic viruses were collected from the Virus Pathogen Database, in addition to additional viruses identified in preclinical models. TREx (Taxonomic-based Relative-Abundance Extractor; https://github.com/TAILOR-Lab/TREx), a companion analysis module for EsViritu, was then used to filter taxonomic profiles based on accession number and taxonomic ID, where non-human viral accessions were first removed. Once filtered, TREx combines each hit to site-level metadata, computing the Reads Per Kilobase per Million Filtered (RPKMF) abundances per sample. This module can aggregate data at the weekly, monthly or yearly level and is publicly available on GitHub^29^.

### Time-Series analysis of average RPKMF of oncogenic viruses

The average RPKMF values calculated with TREx (v0.1) and aggregated monthly across all collection sites were then used as input to plot time-series charts for each oncogenic virus using a custom Python (v3.12.7) script with the matplotlib (v3.9.1) and pandas (v2.3.0) packages. Time-series charts for all viruses were plotted on individual plots as a panel, except for HBV and HCV, where average RPKMF values were plotted within the same plot. To better visualize plots with relatively lower average RPKMF values alongside higher values as a result of plotting multiple viruses on the same plot, dual y-axis plots were generated (**Fig. 3, Supp. Fig. 1**).

### Mapping of reads to Hepatitis A, B, C, D, E references

Reads classified as Hepatitis A, B, C, D and E were mapped against all available reference genomes in the EsViritu Virus Pathogen Database. For each virus, the reference with the highest read support (HAV: LC373510, HBV: AB625342, HCV: NC_004102, HDV: AM779592, HEV: LC055972) was selected for downstream analysis. Coverage was calculated as per-base depth across the full genome, and visualizations included both read depth histograms and feature annotations from corresponding GenBank files. Custom python scripts using pysam (v0.22.1), Biopython (v1.85), and matplotlib (v3.9.1) were used to generate genome-wide coverage plots (**Fig. 2**), with coding regions, including vaccine targets, highlighted for context.

### HPV genome coverage analysis

EsViritu-classified HPV reads were first mapped using a best-to-all strategy to the PaVE database in Geneious Prime (v2025.1.3) using Geneious Assembler with medium sensitivity. For each HPV genus, we the built reference multiple sequence alignments with MAFFT (v7.505) using representative genomes from the PaVE database and selected a single anchor genome per genus (predefined as the genome with the most mapped reads within that genus). We then generated position lift maps from each reference to its genus anchor using the MAFFT alignments as a coordinate guide, and lifted sample BAMs to the common anchor coordinate space with pysam while preserving read-group tags for subtype origin. Per-base depth on the anchor was computed from the lifted BAMs, and coverage summaries were exported as TSV and JSON files. Plot show three alignment panels: coverage across full anchor genome, GenBank-derived features, and read stacks colored by the most contributing types. When indicated, stacked coverage by subtype was rendered from read groups and remaining signal was labeled as “Other.” All scripts (MSA building, lift map creation, BAM lifting, coverage summarization, and plotting) are available and will be released with code repository upon acceptance.

### HPV phylogenetic analysis

Representative HPV genomes were aligned with MAFFT (v7.505) and phylogenetic trees were constructed with IQ-TREE (v2.1.4). Mapped wastewater reads were then used to generate bar graph annotations for each genome, indicating the number of reads mapped. Genus-level rings were also created as annotations for visualization in iTOL. This approach enabled us to place HPV reads within a reference phylogeny, highlight taxonomic breadth across alpha-, beta-, and gamma-papillomaviruses, and annotate clinically relevant types by risk classification and vaccine status.

## Data Availability

Read abundance tables with percent identities of mapping, along with site-specific read counts and taxonomic information will be made publicly available upon manuscript acceptance. While raw genomic sequencing reads cannot be shared due to regulatory considerations, the authors will provide required data to editors and reviewers upon request for peer review purposes.

## Contributors

J.R.C. and A.W.M. conceived the study. Methodology was developed by J.R.C., H.P., R.K.P, M.R., M.T., S.J.J.C., J.D. and A.W.M. Software was designed and implemented by J.R.C., H.P., M.T., and S.J.J.C. Validation and quality control was performed by J.R.C. and H.P. Formal analysis was conducted by J.R.C., H.P. and R.K.P. Investigation was carried out by J.R.C., H.P., R.K.P. Resources were provided by J.R.C. and A.W.M. Data curation was performed by J.R.C., M.T., and S.J.J.C. The original draft was written by J.R.C., H.P. and A.W.M. All authors contributed to manuscript review and editing. Visualization was performed by J.R.C., H.P., R.K.P. and A.W.M. Funding was acquired by E.B. and A.W.M.

## Code Availability

All code developed for read classification, mapping to reference genome, and generation of figures will be made publicly available upon acceptance via dedicated GitHub repository. The authors will provide access to all code upon request for peer review.

## Acknowledgements

Supported by S.B. 1780, 87th Legislature, 2021 Reg. Sess. (Texas 2021) (E.B., A.W.M., and J.F.P.), NIH/NIAID (Grant number U19 AI157981) (A.W.M.), Baylor College of Medicine Joseph Melnick Seed (A.W.M), Alkek Foundation Seed (J.F.P.).

