## Supplemental Figure 1 for "Wastewater Sequencing Reveals Persistent Circulation and Rising Prevalence of Several Oncogenic Viruses Across Texas"

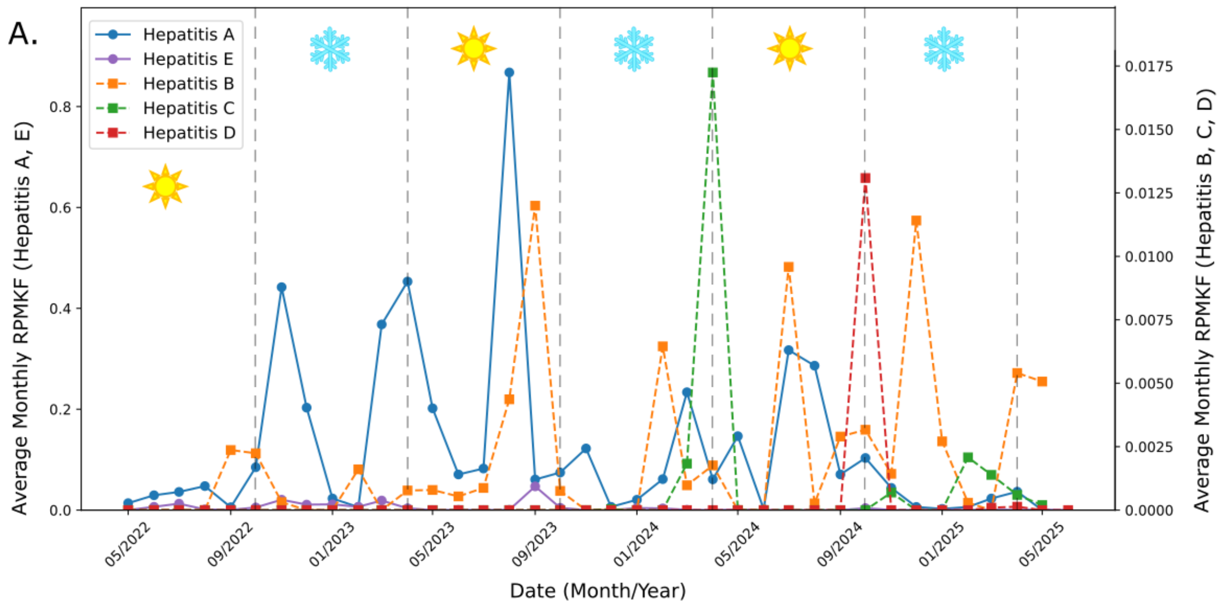


**Supp. Fig 1: Abundance of Hepatitis Family Virus Reads over Three Years of Sampling Across Texas** Mean viral signal of Hepatitis A, B, C, D and E viruses measured in average monthly RPKMF over time in samples taken from wastewater and measure using hybrid-capture probes. Warmer months are highlighted with a sun icon (April-September). Colder months are highlighted with a snowflake icon (October-March). Seasonal boundaries were defined using statewide average temperatures to determine the midpoints between the summer and winter temperature extremes. While Hepatitis A, Hepatitis D and Hepatitis E are not oncogenic themselves, they have been added here to understand Hepatitis transmission dynamics broadly.
